# Prospective sero surveillance among healthcare workers vaccinated with ChAdOx1 nCoV-19 Corona vaccine in a tertiary hospital of Kerala, India

**DOI:** 10.1101/2021.06.29.21259686

**Authors:** Swathi Krishna Njarekkattuvalappil, Ramesh Bhaskaran, Sree Raj V, Ponnu Jose, Aboobacker Mohammed Rafi, Joe Thomas, Susheela J Innah, Lucy Raphael, Unnikrishnan U G, Priyanka Rajmohan, Chithra Valsan, Praveenlal Kuttichira

**Author notes:** **Correspondence:** Ponnu Jose, Assistant Professor, Department of Community Medicine, Jubilee Mission Medical College, Thrissur-680005, Kerala.

## Abstract

**Background:** India started Covid-19 vaccination from January 16, 2021 after the approval of two candidate vaccines namely Covishield ™ and Covaxin ™. We report antibody responses among healthcare workers following two doses of Covishield ™ vaccination in a tertiary care setting.

**Methods:** This prospective serosurveillance study was done among healthcare workers of Jubilee Mission Medical College, vaccinated during January- March 2021. Blood samples were drawn from 170 participants after their 1^st^ dose and from 156 participants after their 2^nd^ dose of Covishield ™ to measure the specific Ig G antibodies against the recombinant S1 subunit of the S protein of SARS-CoV-2.

**Results:** The median level of anti SARS CoV-2 Ig G antibody 28 days after the first dose vaccination is 3.64 S/C (IQR=5.91) and 11.6 S/C (IQR= 5.97) after 14 days of second dose vaccination. Protective levels of anti SARS CoV-2 Ig G antibodies is developed by 25 participants (14.7%) after 28 days of first dose of vaccination and by 109 participants (69.9%) after 14 days of second dose. 18-44 years age group (p=0.027) and absence of comorbidities (p=0.079) are associated with protective IgG levels.

**Conclusions:** Rise in specific Ig G is observed after vaccination. Higher antibody response is observed with younger age group and absence of comorbidities, though statistically not significant. The influence of BMI is also not significant.

## Introduction

The COVID 19 pandemic entered a second peak in India towards early April and is now in a declining mode in all the states including Kerala. The pandemic of Coronavirus disease 2019 (COVID-19) caused by Severe Acute Respiratory Syndrome Coronavirus 2 (SARS-CoV-2) has already affected more than 171 million people and caused more than 3 million deaths worldwide, as of June 04, 2021[1]. To contain this brunt, several novel vaccines recently received an emergency use authorization (EUA) by the U.S Food and Drug Administration (FDA), European Medicine Agency (EMA), U.K. Medicines and Healthcare Products Regulatory Agency (MHRA) and Indian Central Drugs Standard Control Organization (CDSCO) as well as Drugs Controller General of India (DCGI). After receiving EUA, these vaccines were administered to health-care workers, front-line workers, elderly, and at-risk individuals, including people with comorbidities, in a phased roll-out[2].

The vaccination program in India started on January 16, 2021 after the approval of two candidate vaccines namely Covishield ™ (ChAdOx1-nCOV or AZD1222, acquired from Oxford University and AstraZeneca, manufactured by Serum Institute of India, Pune) and Covaxin ™ (BBV-152, manufactured by Bharat Biotech, Hyderabad in collaboration with Indian Council of Medical Research, India)[3].

Our institute also started the vaccination programme with the two dose-regimen of ChAdOx1 nCoV-19 coronavirus vaccine (Covishield™), given 4-6 weeks apart intramuscularly. Covishield™ is a recombinant, replication deficient, chimpanzee adenovirus vector encoding the SARS CoV-2 spike (S) glycoprotein produced in genetically modified human embryonic kidney (HEK) 293 cells. Trials have shown that the vaccine induced a clear antibody response at 28 days after the first dose, across all age groups, including the elderly[4]. As antibody levels correlate with the neutralizing antibody levels; we decided to measure the SARS-CoV-2 Ig G antibody and total antibody to estimate the immune response to the vaccine. The immune response among individuals vaccinated against SARS CoV-2 is hitherto less known. In the light of current vaccination drive by the government with frequently changing dosage intervals, it is imperative to measure the antibody responses to ascertain protection from SARS CoV-2 infection among the individuals. Host factors like age, body mass index, comorbidities also determine the response to vaccines and these have also been examined.

## Methods

Jubilee Mission Medical College & Research Institute is a 1600 bedded teaching hospital with around 3000 staff in the regular pay rolls, daily wagers, and workers of service contractors. The institute started vaccination of its staff with ChAdOx1 nCoV-19 coronavirus vaccine, as per the Government of India guidelines, using the vaccine supplied by the Kerala Health Services on 19 January 2021. Baseline antibody levels of individuals against SARS CoV-2, prior to vaccination, is available with us as part of the study conducted in our institute during September to December 2020[5]. The current study is a prospective sero surveillance study initiated after obtaining the Institutional Ethics Committee approval from January 2021 to April 2021.

Participants of the study were healthcare workers of JMMC & RI: (a) who have provided their pre-vaccination serum sample for SARS CoV-2 antibody estimation and found negative (b) who have taken the first dose of ChAdOx1 nCoV-19 Coronavirus vaccine (c) with no history or test result suggestive of Covid infection. Satisfaction of all the three criteria was essential to be part of the study. After obtaining informed consent, a self-administered questionnaire in Google forms was filled by the participant (Annexure 1). On the day of the second dose of vaccination, which ranged from 28 to 56 days after the first dose, a 5 ml blood sample was drawn by a trained phlebotomy team. Similarly, blood samples were also drawn 14 days after the 2^nd^ dose (i.e. 42 days after 1^st^ dose) from the same set of participants. A total of 170 individuals provided their blood samples after the first dose of vaccination. 14 participants were excluded from the second analysis of the study as they refused to provide consent for repeat blood samples. The blood samples collected were centrifuged and plasma separated and frozen at -20° Celsius for batch testing. The samples were subjected to SARS-CoV-2 Ig G and total antibody testing using the VITROS anti SARS CoV-2 Ig G/Total Chemiluminescence kit manufactured by Ortho Clinical Diagnostics, USA (OCD). Both the VITROS anti-SARS-CoV-2 Total and Ig G assays (Ortho Clinical Diagnostics) are based on CLIA using luminol-horseradish peroxidase (HRP)-mediated chemiluminescence. Both assays were performed on the VITROS 3600 automated immunoassay analyzer (Ortho Clinical Diagnostics) according to the manufacturer’s instructions. In these assays, the specific antibodies against the recombinant S1 subunit of the S protein of SARS-CoV-2 were automatically analyzed. Results are reported as signal/cut-off (S/C) values and as qualitative results indicating non-reactive (S/C < 1.0; negative) or reactive (S/C ≥ 1.0; positive). The VITROS anti-SARS-CoV-2 total assay can detect total antibodies (IgA, Ig M, and Ig G) against SARS-CoV-2 S protein. Anti SARS Cov-2 Ig G antibody levels have been tested using various platforms especially for the extraction of high titre Covid-19 Convalescent Plasma. The protective levels post vaccination has not yet been validated. The protective level for convalescent plasma is considered as above 9.5 S/C as per the US FDA document published for use in the manufacture of high titre Covid-19 Convalescent Plasma [6].

Data collected was entered into Microsoft Excel spread sheets and analyzed using IBM Statistical Package for Social Sciences (SPSS) version 25. Categorical variables are expressed as proportions/percentages; continuous variables as mean/median with standard deviation/interquartile range and association of various factors with immune response are estimated by regression method.

## RESULTS

The study was conducted among 170 healthcare workers of different cadres who received Covishield ™ vaccination. Table 1 describes baseline characteristics of the population. The mean age of the study population is 37.09 years (SD=12.6). 126 (74.1%) belong to the 18-44 years age group, 30 (17.6%) to the 45-59 years age group and 14 (8.2%) to the 60 years and above age group. Males constituted 23.5% and females 76.5%. The body mass index (BMI) categories according to the Asian classification[7] show that 21 (12.4%) are underweight, 45 (26.5%) normal, 40 (23.5%) overweight and 64 (37.6%) obese. 23(19.4%) participants have comorbidities like diabetes, hypertension, bronchial asthma etc. 144 (84.7%) have a previous history of BCG vaccination and 17 (10%) have a history of influenza vaccination.

**Table 1:** Characteristics of study population (n=170)

|  |  |  |
| --- | --- | --- |
| **Age** |  |  |
|  | Mean (SD) | 37.09 years (12.6) |
|  | Median (IQR) | 34 years (17) |
|  | 18-44 years | 126 (74.1%) |
|  | 45-59 years | 30 (17.6%) |
|  | ≥60 years | 14 (8.2%) |
| **Gender** |  |  |
|  | Male | 40 (23.5%) |
|  | Female | 130 (76.5%) |
| **Body Mass Index (BMI)** |  |  |
|  | Underweight | 21 (12.4%) |
|  | Normal | 45 (26.5%) |
|  | Overweight | 40 (23.5%) |
|  | Obese | 64 (37.6%) |
| **Comorbidity** |  | 33 (19.4%) |
| **Previous BCG vaccination** |  |  |
|  | Yes | 144 (84.7%) |
|  | No | 12 (7.1%) |
|  | I don't know | 14 (8.2%) |
| **Previous influenza vaccination** |  |  |
|  | Yes | 17 (10%) |
|  | No | 115 (67.7%) |
|  | I do not know | 38 (22.4%) |
| **Last vaccine received** |  |  |
|  | During childhood | 24 (14.1%) |
|  | More than 10 years ago | 31 (18.2%) |
|  | 5 to 10 years ago | 35 (20.6%) |
|  | Within last 5 years | 80 (47.1%) |

Adverse events following vaccination are reported by 141 participants. Fever (88/170) is the commonest symptom, followed by pain at the injection site (59/170), myalgia (59/170) and headache (45/170).

Protective levels of anti SARS CoV-2 Ig G antibodies are observed in 14.7% (n=170) participants after the first dose and in 69.9% (n=156) individuals after the second dose. Table 2 describes the antibody levels at different time points. The median level of anti SARS CoV-2 Ig G antibody in the serum of 170 participants, 28 days after the first dose vaccination, is 3.64 S/C (IQR=5.91). Figure 1 depicts the distribution of serum antibodies 28 days and 42 days after first dose of vaccine. The median antibody level after 14 days of second dose vaccination for 156 participants is 11.6 S/C (IQR= 5.97).

**Table 2:** Levels of anti SARS CoV-2 Ig G antibodies in individuals vaccinated with Covishield.

|  |  |  |
| --- | --- | --- |
| **After 28 days of first dose (n=170)** |  |  |
| Protective levels ($\geq 9.5$ S/C) | | 25 (14.7%) |
| Less than 9.5 S/C |  | 145 (85.3%) |
| **After 14 days of second dose (n=156)** |  |  |
| Protective levels ($\geq 9.5$ S/C) | | 109 (69.9%) |
| Less than 9.5 S/C |  | 47 (30.1%) |

**Figure 1:**
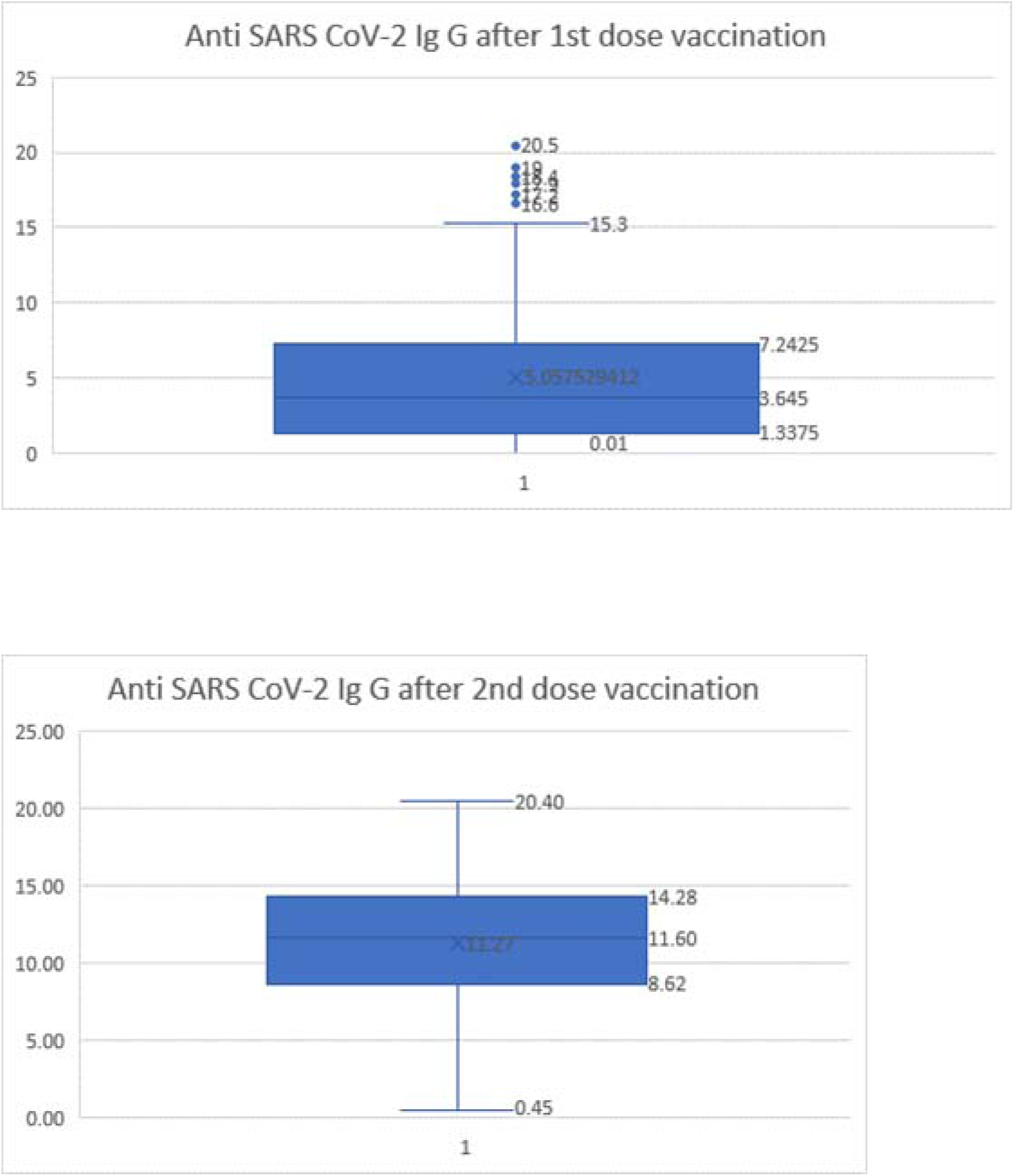
Anti SARS Cov-2 Ig G antibody levels in individuals 28 days and 42 days after first dose of Covishield vaccine.

**Figure 2:**
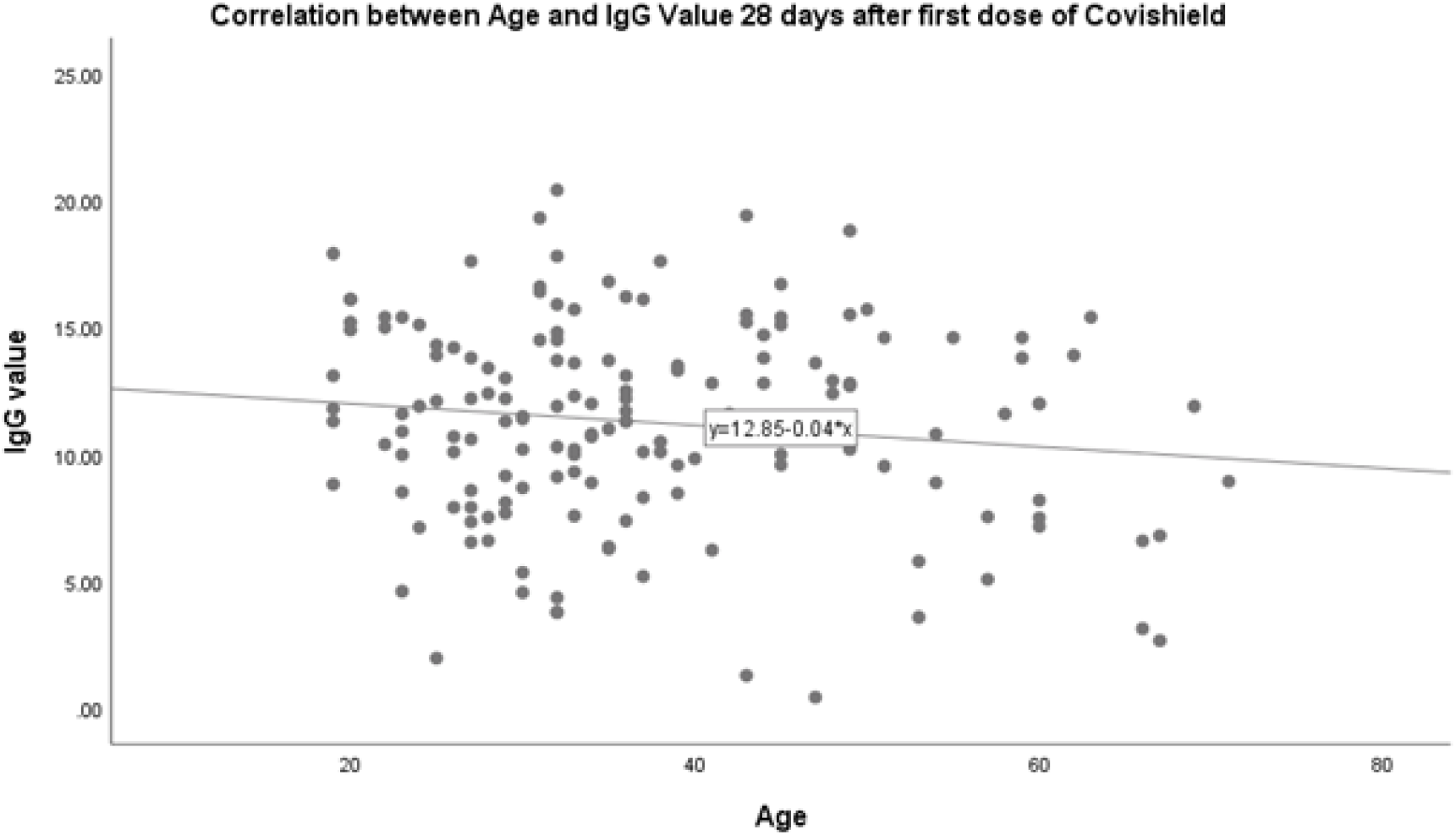
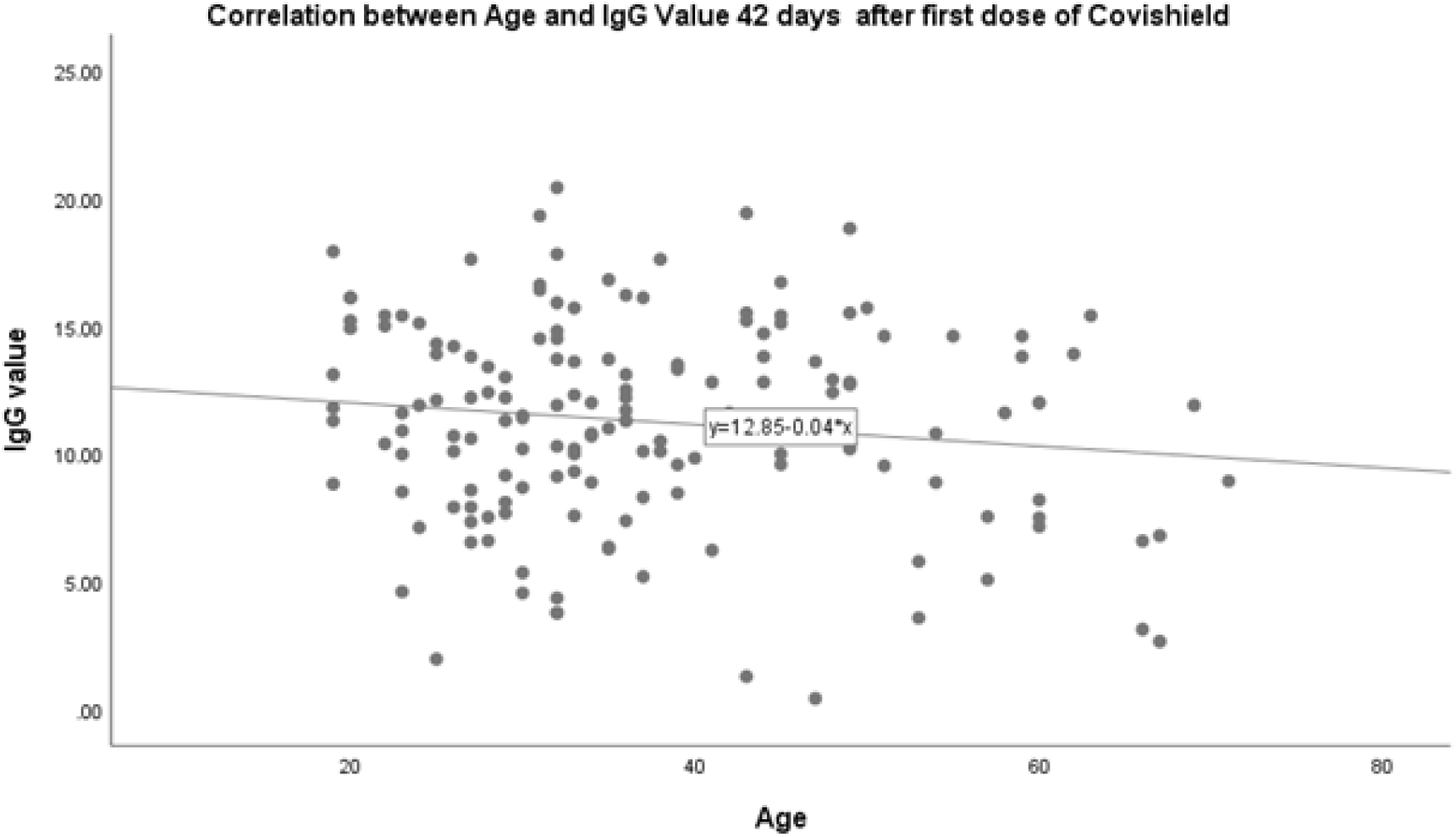
Scatter plot showing relationship between age and serum Ig G levels of anti SARS CoV-2 Ig G antibodies 28 days (r= -0.19) and 42 days after first dose of Covishield (r=-0.13)

Sub-group analysis (Table 3) of the participants who developed protective levels of Ig G following vaccination revealed that 80% of the participants after the first dose and 75.2% of participants following the second dose belong to the 18-44 years age group.

**Table 3:** Profile and association of individuals who developed protective levels of Ig G (≥ 9.5 S/C) after Covishield vaccination

| Characteristic | Participants with protective antibody levels after first dose (25/170) | p Value | Participants with protective antibody levels after second dose (109/156) | p Value |
| --- | --- | --- | --- | --- |
| **Age** |  |  |  |  |
| Mean (SD) | 33.04 years (12.6) |  | 36.32 years (11.4) |  |
| Median (IQR) | 32 years (19.5) |  | 34 years (17) |  |
| 18-44 years | 20 (80%) |  | 82 (75.2%) |  |
| 45-59 years | 4 (16%) | 0.664 | 22 (20.2%) | 0.027 |
| > or = 60 years | 1 (4%) |  | 5 (4.6%) |  |
| **Gender** |  |  |  |  |
| Male | 3 (12%) |  | 27 (24.8%) |  |
| Female | 22 (88%) | 0.141 | 82 (75.2%) | 0.725 |
| **Body Mass Index** |  |  |  |  |
| Underweight | 4 (16%) |  | 12 (11%) |  |
| Normal | 3 (12%) | 0.324 | 33 (30.3%) | 0.667 |
| Overweight | 6 (24%) |  | 25 (22.9%) |  |
| Obese | 12 (48%) |  | 39 (35.8%) |  |
| **Comorbidity** |  |  |  |  |
| Yes | 5 (25.0%) | 0.936 | 17 (15.6%) | 0.079 |
| No | 20 (75.0%) |  | 92 (84.4%) |  |
| **Interval between two doses of vaccination** |  |  |  |  |
| > 42 days (n=33) | - | NA | 23 (21.1%) | 0.980 |
| ≤ 42 days (n=123) | - |  | 86 (78.9%) |  |

Independent analysis of various factors for association with serum Ig G levels of anti-SARS CoV-2 antibodies after second dose vaccination. Tables (3) showed significant association of younger age groups (below 60 years age) (p=0.027) in building up protective antibody levels. Logistic regression analysis of these factors showed no significant association. We did linear regression of age of the individual as well as time interval between two doses with antibody levels after first and second doses of vaccination, which revealed no correlation. The Pearson correlation coefficient (r) for age and time gap with serum Ig G levels and scatter plots are given in Figures (2).(3)

**Figure 3:**
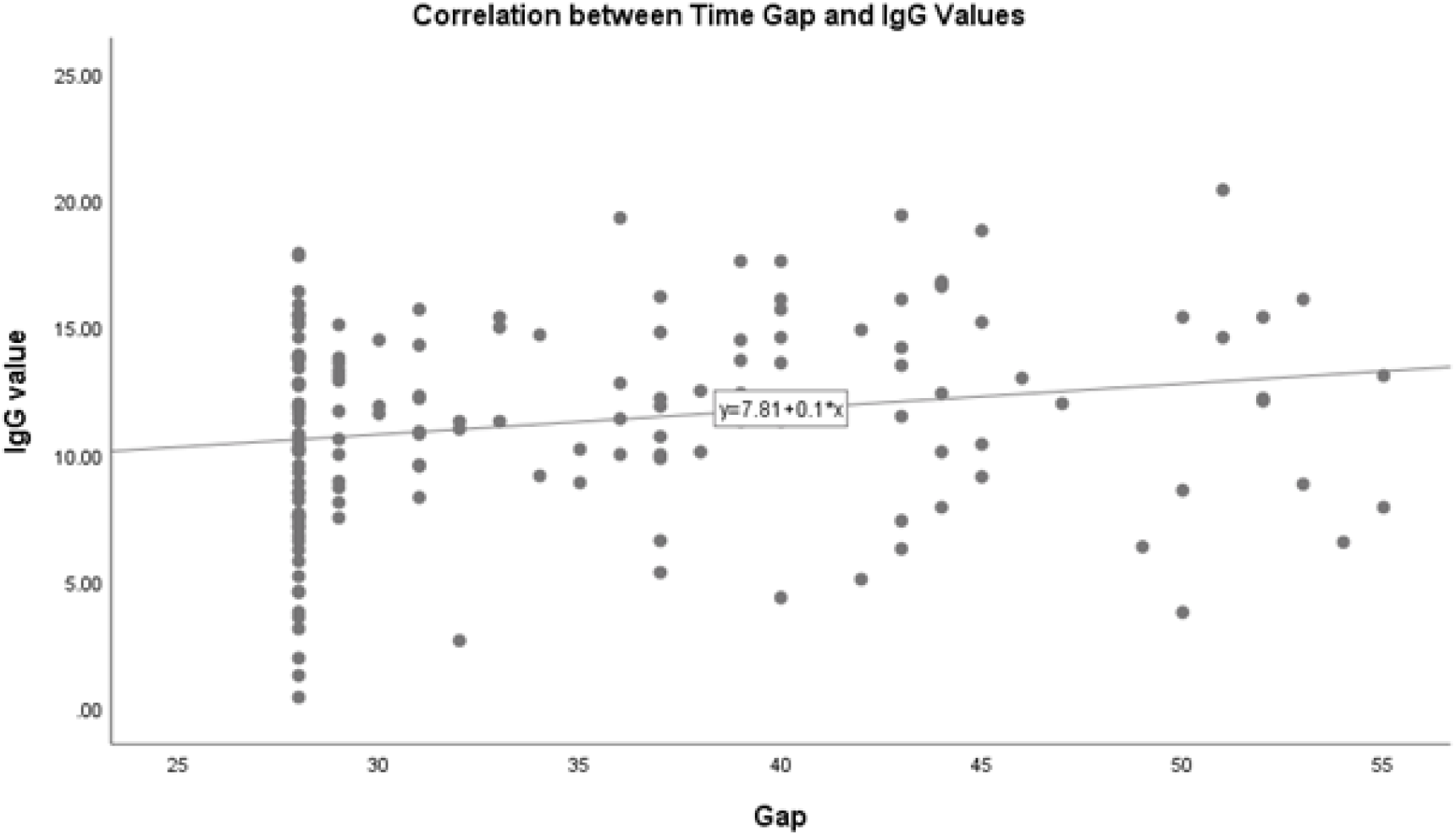
Scatter plot showing relationship between time gap between two doses of Covishield and serum Ig G levels of anti SARS CoV-2 Ig G antibodies (r = 0.19)

## Discussion

Antibody response plays a major role in developing protective immunity during SARS-CoV-2 infection [8]. Although sero-positivity based on different cut off values of I g G titres does not necessarily translate into direct protection from SARS-CoV-2 infection, it can be used as a surrogate biomarker of immunity in assessing the human humoral response to Covid-19. In this study, we assessed the humoral immune response after the first and second doses of Covishield™ vaccine in healthcare workers (HCWs) in a tertiary hospital in Kerala. The samples were subjected to SARS-CoV-2 Ig G and total antibody testing using the VITROS anti SARS CoV-2 Ig G/Total Chemi luminescence kit manufactured by Ortho Clinical Diagnostics, USA, which has already been validated to have a high specificity in detection of antibodies in convalescent plasma of Covid infected patients [9].However, the protective levels of antibody, post vaccination has not yet been validated. Hence we followed the cut-off value of 9.5 S/C as prescribed by the manufacturer based on protective levels required for convalescent plasma [6]. It has already been proved in various studies[10],[11] that Ig G levels following natural infection with SARS CoV-2 can persist for several months, including an ongoing study on a cohort of Belgian hospital workers, which shows persistence of Ig G till upto 199 days[12]. This principle may also be applied for antibody response following vaccine administration.

From a pooled analysis of four randomized trials on immunogenicity and efficacy of ChAdOx1 nCoV-19 (AZD1222) vaccine by Voysey et al. it is already known that individuals aged 18–55 years who received a second dose vaccine more than 12 weeks after the first had antibody titres more than two-fold higher than those who received the second dose within 6 weeks of their initial vaccination [4]. In our study, among the 170 HCWs, protective antibody levels were found in 14.7% after first dose which rose to 69.9 % after the second dose of Covishield™ vaccine. Katie J Ewer et al reports an increase of spike specific IgG activity between day 28 and 56 after dosing [13] and Folegatti et al reports an increase in protective neutralizing antibody levels after second dose [14]. Our observations are in concordance with these two studies. However, an Indian study by Awadhesh et al among HCWs reports 79.3% positivity which is higher than our findings [15]. Inclusion of subjects with history of Covid-19 could be a reason to explain the faster high antibody level in this report. We excluded from our study, participants with a history of Covid-19 infection. Qadri et al from the International Centre for Diarrhoeal Disease Research, Bangladesh reports that more than 90% of study subjects developed antibodies 30 days after Oxford-AstraZeneca Covid-19 vaccination which rose to 97% after two months[16]. Prior infection with Covid-19 shows a fourfold increase in the levels. Study conducted in Pakistan by Umar et al using Gam-Covid-Vac (Sputnik V, Russian made) found 85% of study subjects developing SARS-CoV-2 spike protein antibodies, with higher levels for those having previous infection history[17].

Subgroup analysis of those participants who built up protective antibody levels in our study reveals a preponderance of 18-44 years age group (p=0.027). 80% participants who developed Ig G more than 9.5 S/C following first dose vaccination and 75.2% following second dose belong to this particular age group. The majority of above 60 years age group remained to be ‘low-responders’ despite two doses of vaccination. Similar findings are reported in age-dependent immune response studies to the BioNTech/Pfizer BNT162b2 Covid-19 vaccine and AstraZeneca vaccine by Gareth et al. and Wei et al[18],[19].The study by Kataria S et al in the National Capital Region of India finds a significantly higher titres of SARS CoV-2 S1/S2 IgG in age less than 50 years [20]. Perhaps aging and resultant immunosenescence may have a role in the decreased response to vaccinations in the elderly population[21],[22].

In the current study, even though 73% of participants without comorbidities like diabetes, hypertension and bronchial asthma developed protective levels of antibody, it is not significant statistically (p=0.079). Literature shows no differences in immune responses with presence or absence of comorbidities following natural infection with the virus [23]. The limited studies available following vaccination also show no significant association of comorbidities with antibody response[16].

The time interval between two doses of the vaccine in our study ranges from 28 to 56 days. Even though there is enough evidence to show that an increasing time interval between two doses of Covishield™ is beneficial in building up antibody response[24], we could not observe any such significant association (figure 3). This might be a function of the small sample size of our study.

This is a prospective serosurveillance study following up a cohort of vaccinated healthcare workers for one year. These are the preliminary results of the study. Our limitations include a small sample size and exclusion of individuals who developed Covid-19 infection before or in between vaccination. We have also not looked into cell-mediated immune responses which could augment responses against Covid-19 virus.

## Conclusion

Evidence base for optimal vaccine dosage, timings of doses, adverse events following immunization and need for booster dose remains scarce for Covid 19 vaccinations. The current study observed positive antibody response to the first dose of Covishield vaccine which is enhanced after the second dose. Antibody response in younger age groups is higher and the influence of BMI and co-morbidity is not significant. Given the current scenario in India where there is scarcity of hospital beds, oxygen and ventilators coupled with low supply of vaccines, an effective strategy for vaccination is needed. Our study results provide a baseline data and supplement the evidence for the same.

## Data Availability

data will be available on request

## LIST OF ABBREVIATIONS

SARS: CoV-2 Severe Acute Respiratory Virus Coronavirus- type 2
HCW: Health Care Worker
AEFI: Adverse Events Following Immunization
Ig G: Immunoglobulin Gamma

## DECLARATIONS

### Ethics approval and consent to participate

The study protocol has been approved by the Institutional Ethic Committee of Jubilee Mission Medical College & Research Institute, Thrissur (IEC study ref no: 38/21/IEC/JMMC & RI dated 18-02-2021).

### Consent for publication

Written informed consent was obtained from each participant prior to enrolment and blood sample draw.

### Availability of data and material

Data collected was compiled into excel sheets by SRV & AMR under the supervision of SJI. Data cleaning, analysis and review was done by SKN, UUG and PJ.

### Authors’ contributions

SKN, AMR and PJ conceived and designed the study and developed the study protocol and analysis plan. SJI and JT coordinated the project. SKN, AMR, SRV and PJ coordinated data collection at the site. SKN, UUG and PJ analyzed the data. SKN and RB drafted the manuscript. All authors have reviewed the manuscript and approved it. PK, SJI and JT had complete access to the data and guarantee the manuscript.

## Acknowledgements

The authors would like to acknowledge the efforts of Dr Mimitha A M, Mr Anson Abraham, Mr Manoharan, Ms Manju, phlebotomists & technical staff from the central lab for the coordination, sample collection at the vaccination centre, sample processing and storage.

